# Thalamohippocampal atrophy in focal epilepsy of unknown cause at the time of diagnosis

**DOI:** 10.1101/2020.07.24.20159368

**Authors:** Nicola J. Leek, Barbara A. K. Kreilkamp, Mollie Neason, Christophe de Bezenac, Besa Ziso, Samia Elkommos, Kumar Das, Anthony G. Marson, Simon S. Keller

**Affiliations:** Department of Pharmacology and Therapeutics, Institute of Systems, Molecular and Integrative Biology, University of Liverpool, UK; The Walton Centre NHS Foundation Trust, Liverpool, UK; St George’s University Hospitals NHS Foundation Trust, London, UK

**Keywords:** Newly diagnosed epilepsy, focal epilepsy, thalamus, hippocampus, treatment outcome

## Abstract

**Background:** Patients with chronic focal epilepsy may have atrophy of brain structures important for the generation and maintenance of seizures. However, little research has been conducted in patients with newly diagnosed focal epilepsy (NDfE), despite it being a crucial point in time for understanding the underlying biology of the disorder. We aimed to determine whether patients with NDfE show evidence of volumetric abnormalities of subcortical structures.

**Methods:** Eighty-two patients with NDfE and 40 healthy controls underwent MRI scanning using a standard clinical protocol. Volume estimation of the left and right hippocampus, thalamus, caudate nucleus, putamen and cerebral hemisphere was performed for all participants and normalised to whole brain volume. Volumes lower than two standard deviations below the control mean were considered abnormal. Volumes were analysed with respect to patient clinical characteristics, including treatment outcome 12 months after diagnosis.

**Results:** Volume of the left hippocampus (*P*_(FDR-corr)_ = 0.04) and left (*P*_(FDR-corr)_ = 0.002) and right (*P*_(FDR-corr)_ = 0.04) thalamus were significantly smaller in patients relative to controls. Relative to the normal volume limits in controls, 11% individual patients had left hippocampal atrophy, 17% had left thalamic atrophy and 9% had right thalamic atrophy. We did not find evidence of a relationship between volumes and future seizure control or with other clinical characteristics of epilepsy.

**Conclusions:** Volumetric abnormalities of structures known to be important for the generation and maintenance of focal seizures are established at the time of epilepsy diagnosis and are not necessarily a result of the chronicity of the disorder.

## Introduction

There is a wealth of evidence indicating that people with refractory focal epilepsy have quantitative structural brain abnormalities on magnetic resonance imaging (MRI). Atrophy of temporal lobe structures is frequently identified in patients with temporal lobe epilepsy (TLE), including the hippocampus, entorhinal cortex, perirhinal cortex and amygdala, preferentially ipsilateral to the side of seizure onset ^[1]^. Extrahippocampal subcortical atrophy is also commonly reported, including the thalamus and striatum in both cerebral hemispheres ^[1, 2]^. In TLE seizures are primarily generated in the hippocampus; in vitro electrophysiology of resected hippocampal tissue from refractory patients has shown the dentate gyrus to be the most susceptible region to the generation of epileptiform activity ^[3]^. Intracerebral recordings of electroencephalography (EEG) activity in patients with refractory focal epilepsy exhibit an increase of synchrony between the thalamus and temporal lobe structures during seizures ^[4]^. Furthermore, an increase in neuronal firing rate in the caudate nucleus and putamen has been observed during prefrontal seizures in an animal model of epilepsy ^[5]^. Therefore, there is a well-established link between subcortical atrophy and brain seizure activity in patients with chronic focal epilepsy. However, it remains unclear whether subcortical atrophy in focal epilepsy is pre-existing, present at the time of diagnosis as a consequence of epileptogenic processes, or the result of the chronicity of longstanding epilepsy and antiepileptic drug (AED) treatment. It is therefore important to determine whether brain abnormalities are already established in the early stages of epilepsy.

Despite that epileptogenesis begins prior to the onset of a first seizure ^[6]^, the earliest reliable time point of investigation of human epilepsy in prospective studies is at the point of diagnosis. Neuroimaging studies of patients with newly diagnosed epilepsy (NDE) have the potential to provide important information about the nature of brain abnormalities by separating pre-existing or novel abnormalities and longstanding changes originating from recurrent seizures and chronic use of AEDs ^[7]^. The identification of quantitative imaging abnormalities at diagnosis may provide new insights into biomarkers of pharmacoresistance and cognitive comorbidities ^[8, 9]^. Approximately 60% of patients with NDE will achieve seizure control, ∼25% will develop pharmacoresistant epilepsy and the remainder will fluctuate between remission and relapse ^[10]^. To date, markers of pharmacoresistance in patients with NDE have been limited to reports in epidemiological studies and clinical trials, and suggest, for example, that gender, treatment history, age, and time between first seizure and diagnosis may be related to pharmacoresistance ^[11, 12]^. Determining the relationship between quantitative brain imaging at diagnosis and AED treatment outcome is an important research endeavour ^[8]^. Additionally, over 50% of patients with NDE have been found to show impairment in at least one cognitive domain ^[13]^. Quantitative imaging studies may further contribute to the understanding of cognitive problems which may be present at point of diagnosis.

There are few quantitative neuroimaging studies in patients with newly diagnosed focal epilepsy (NDfE). Although a small number of studies have identified localised brain atrophy in patients with NDfE, findings are inconsistent. One study revealed hippocampal atrophy in patients with NDfE relative to controls ^[14]^, whilst others have found no difference in hippocampal volume between these groups ^[15, 16]^. Inconsistent findings have been reported on structural changes in the cerebellum in patients with NDfE ^[16, 17]^. In a small-scale study using voxel-based morphometry, we were unable to identify morphometric alterations of subcortical or cortical regions in patients with NDfE compared to controls ^[18]^. Although, to our knowledge, no studies have identified thalamic atrophy in adult NDfE, a recent study reported thalamic atrophy in drug naïve patients with new-onset genetic generalised epilepsy (GGE) ^[19]^.

There were two primary objectives of the present study. Firstly, we sought to determine whether atrophy of the hippocampus, thalamus, caudate nucleus, putamen and cerebral hemisphere is present at the time of diagnosis of focal epilepsy with unknown cause relative to healthy controls. Secondly, we aimed to explore whether volumetric changes of these structures are related to various clinical characteristics of the disorder including treatment outcome at 6 and 12 months after diagnosis.

## Methods

### Participants

We identified patients with archived MRI, acquired according to a clinical epilepsy protocol, within 12 months of diagnosis of focal epilepsy of unknown cause and scanned on a 3 T GE Discovery MRI system at the Walton Centre NHS Foundation Trust, Liverpool, UK since 2015. At initial screening, 140 patients with likely NDfE and corresponding MRI for analysis were retrieved. More detailed assessment of patient clinical histories and MRI resulted in the exclusion of 58 patients due to one of the following factors: (1) first seizure with no diagnosis of epilepsy, (2) probable idiopathic generalised epilepsy, (3) symptomatic seizures (e.g. tumour, infection), (4) presence of epileptogenic lesion (e.g. focal cortical dysplasia, hippocampal sclerosis), or (5) unusable or unavailable MRI data for analysis. This resulted in 82 patients with NDfE of unknown cause with corresponding MRI data for image analysis (Table 1). All patients were diagnosed by consultant neurologists at the Walton Centre NHS Foundation Trust. All images were reported non-lesional by a neuroradiologist with expertise in the assessment of MRI for epileptogenic lesions. All patients had no history of learning disability.

**Table 1.** Demographic and clinical data. Mean age at diagnosis / MRI is presented with standard deviation. MRI, magnetic resonance imaging; FAS, focal aware seizures; FIAS, focal impaired awareness seizures; FBTCS, focal to bilateral tonic-clonic seizures; EEG, electroencephalography. *Significantly different (_*X*2_ = 4.72, *P* = 0.03).

| Clinical variable | Patients | Controls |
| --- | --- | --- |
| *n* | 82 | 40 |
| Age at diagnosis / MRI | 38.04 (10.8) | 32.50 (8.9) |
| Sex | 50 M / 32 F* | 16 M / 24 F* |
| FAS | 35 / 82 (42.7%) | - |
| FIAS | 47 / 82 (57.3%) | - |
| FBTCS | 72 / 82 (87.8%) | - |
| Normal EEG | 39 / 51 (76.5%) | - |
| Abnormal EEG | 12 / 51 (23.5%) | - |
| Seizure free - 6 months | 23 / 58 (39.7%) | - |
| Seizure free - 12 months | 24 / 48 (50%) | - |

Additional clinical data were obtained by searching through hospital electronic records. We obtained age at diagnosis and seizure type (focal aware seizures [FAS], focal impaired awareness seizures [FIAS] and focal to bilateral tonic-clonic seizures [FBTCS]) for patients. Fifty-one (62.2%) had undergone EEG and we recorded whether inter-ictal abnormalities were captured. Seizure status at 6 and 12 months after diagnosis, hereon referred to as seizure outcome, was obtained for 58 patients at 6 months and 48 patients at 12 months.

For comparison with patients, we used imaging data from a cohort of 40 healthy adult controls that were scanned as part of a different study ^[20]^ but who had the equivalent MRI scans for comparative analysis (Table 1). There was no significant difference between the age of patients (when diagnosed) and controls (at time of MRI) (*t* = 2.8, *P* = 0.13). However, there was a significant sex difference between patients and controls (*X*^2^ = 4.72, *P* = 0.03). The North West – Liverpool research ethics committee approved this study (14/NW/0332).

### MRI acquisition

The standard 3 Tesla MRI protocol for patients with a new presentation of seizures at our centre included a high in-plane resolution T1-weighted fluid attenuated inversion recovery (FLAIR) MRI acquisition of the whole brain (TE 1.5 ms, TR 2500 ms, flip angle 111°, voxel size 0.4 mm × 0.4 mm, slice thickness 3.0 mm, FOV 220 mm, matrix size 320 × 384). This sequence was used for analysis in the present study. Other sequences acquired for diagnostic purposes but not used for analysis included axial T2-weighted and coronal T2-FLAIR scans.

### MRI analysis

Given the slice thickness of the T1-weighted FLAIR images, we were unable to reliably apply automated image analysis tools to extract subcortical and hemispheric volume. We therefore used rigorous manual techniques to estimate the volume of subcortical and hemispheric structures. The volume of the left and right hippocampus, thalamus, caudate nucleus, putamen, and cerebral hemispheres were quantified for all participants using the Cavalieri method of design-based stereology ^[21]^. This approach has been frequently applied to MRI data in epilepsy studies ^[15, 22-24]^, has been considered the benchmark measurement approach to which automated MRI techniques have been compared ^[23]^, and provides a mathematically unbiased and validated approach to estimate brain compartment volume ^[21, 25]^.

Using Easymeasure software ^[23]^, each volume of interest (VOI) was estimated using a series of parallel two-dimensional (2D) MR sections set at a constant distance apart. A randomly orientated grid of pixels was overlaid on each section and points intersecting each region-of-interest (ROI) were counted separately for the left and right structures of each patient and control. The pixel size used for point-counting was altered depending on the size of the ROI (pixel sizes: hippocampus, 4; thalamus, 6; caudate nucleus, 5; putamen, 6; and cerebral hemisphere, 30) in order to optimise the sampling density ^[21]^. The number of points transecting the ROI was multiplied by distance between each consecutive section to produce volume estimates. Given that nuclei (e.g. thalamic nuclei) and subregions (e.g. hippocampal cornu ammonis) of structures measured are almost indistinguishable on clinical MRI, we measured the structures as an entire complex.

Stereological point counting on MR images for volume estimation of the hippocampus, thalamus, caudate nucleus and putamen is shown in Figure 1. Detailed information on the hippocampal VOI is provided elsewhere ^[24]^. Moving along its longitudinal axis, the hippocampus is bound superiorly by the white, myelinated fibres of the alveus and often by an additional region of cerebrospinal fluid superior to the alveus. The hippocampus was differentiated anteriorly from the amygdala through visualisation of the alveus. The posterior boundary of the hippocampus was reached when the lateral ventricles divide into the frontal and temporal horns. The hippocampal VOI comprised the hippocampus proper, dentate gyrus, alveus, subiculum, presubiculum and parasubiculum; the amygdala, uncus, choroid plexus and grey matter above the alveus were not included in the measurements.

**Figure 1.**
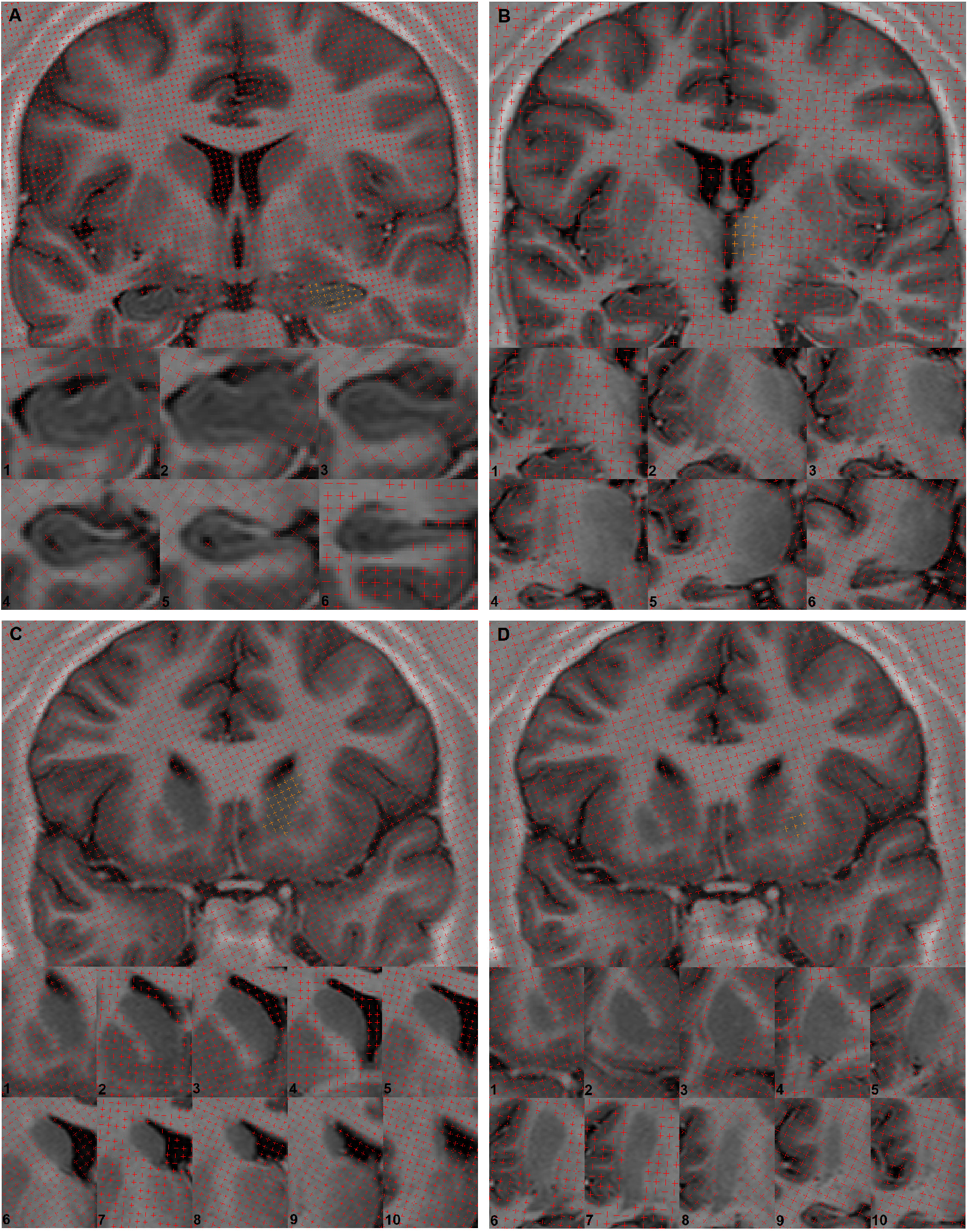
T1-weighted fluid attenuated inversion recovery coronal sections through an exemplar patient showing point counting for stereology through the subcortical volumes of interest. For each structure, point counts are removed in the left hemisphere and coloured orange in the right hemisphere. A, hippocampus; B, thalamus; C, caudate nucleus; D, putamen. Zoomed sections at the bottom of each panel show point counting in a rostral (top left) to caudal (bottom right) direction.

The anterior border of the thalamus began immediately posterior to the anterior commissure and maintained a close relationship with the internal capsule laterally and the central canal of the ventricles medially; the posterior border of the thalamus was the pulvinar. Measurements ended with the formation of the atrium of the ventricles. The zona incerta formed the inferior border of the thalamus. We excluded the subthalamic nuclei, substantia nigra, and red nuclei from thalamic measurements. The lateral and medial geniculate bodies and the habenular nucleus were also excluded ^[23]^. The posterior border of the caudate nucleus was considered the last slide in which the caudate tail was still superior to the lateral ventricle. Caudate nucleus and putamen measurements ended with the formation of the atrium from the temporal and frontal horns of the lateral ventricle. Neither caudate nucleus nor putamen measurements included the striatal cell bridges connecting the two nuclei or the nucleus accumbens. The medial and lateral borders of the putamen were the internal capsule and external capsule, respectively ^[22]^. The posterior border of the putamen often coincides with the appearance of the medial and lateral geniculate bodies. Measurement of the entire cerebral hemispheres was also obtained which included all supratentorial grey and white matter, excluding the brainstem and cerebellum. All subcortical and hemispheric volumes were normalised using whole brain volumes (summation of left and right cerebral hemispheric volumes); the proportion of each subcortical and hemispheric volume relative to whole brain volume was calculated.

### Statistical analysis

The data did not meet the assumptions of parametric tests; therefore, nonparametric tests were used to analyse the data. For patient-control analysis, all volumes were analysed using nonparametric Mann-Whitney U-tests in SPSS (version 25, www.spss.com). Given the significant sex difference between patients and controls, and the higher average age of patients compared to controls, we performed all statistical analyses on normalised volumes, with their residuals corrected for age and sex in a confound only regression model. Volumes lower than two standard deviations of the control mean were considered abnormal and suggestive of structural atrophy in individual patients. Spearman’s correlations were used to investigate relationships between volumes and age at first seizure, years between first seizure and diagnosis, and age at diagnosis. Categorical analysis of clinical and neuroimaging data was performed using Chi-squared tests. Multiple comparisons were corrected using the false discovery rate (FDR) and results were considered statistically significant at *P* < 0.05.

## Results

### Clinical data

Two patients did not commence AED treatment. These patients did not have outcome data. For the remainder of the patient cohort, the first AED used was Lamotrigine (*n* = 47, 59%), Levetiracetam (*n* = 14, 18%), Zonisamide (*n* = 8, 10%), Carbamazepine (*n* = 6, 8%), Sodium Valproate (*n* = 3, 4%), Oxcarbazepine (*n* = 1, 1%) and Phenytoin (*n* = 1, 1%). There were no significant associations in the clinical data.

### Volumetric changes in NDfE

Volumetric descriptive statistics are provided in Table 2. Volume of the left hippocampus (*U* = 1232, *P*_(FDR-corr)_ = 0.04), and left (*U* = 1002, *P*_(FDR-corr)_ = 0.002) and right (*U* = 1228, *P*_(FDR-corr)_ = 0.04) thalamus were significantly smaller in patients compared to controls (Figure 2). There was a trend for the right hippocampus to be smaller in patients relative to controls (*U* = 1311, *P*_(FDR-corr)_ = 0.09). There were no significant differences (*P*_(FDR-corr)_ < 0.05) or trends for differences in volume of the left or right caudate nucleus, putamen or whole cerebral hemisphere between patients and controls. In patients, average volume of the left hippocampus was decreased by 6.9%, right hippocampus by 6.5%, left thalamus by 6.4%, and right thalamus by 4.1%, relative to controls. Individual volumetric analysis revealed abnormal volume of the left hippocampus in 9 (11%) patients, the right hippocampus in 4 (4.9%) patients, the left thalamus in 14 (17.1%) patients, and the right thalamus in 7 (8.5%) patients.

**Table 2.** Results of volumetric comparisons between patients and controls. Mean and standard deviation (SD) of subcortical and hemispheric volumes, expressed as percentage of whole brain volume and their residuals corrected for age and sex (%CorrAgeSex), and raw volume (cm^3^). For each structure the number (and percentage) of patients with volumes lower than the normal limits is indicated as abnormal (*n*, %).

| Structure | Patients |  |  | Controls |  |  | *U, P(FDRcorr)* |
| --- | --- | --- | --- | --- | --- | --- | --- |
|  | Mean | SD | Abnormal<br>(*n, %*) | Mean | SD | Abnormal<br>(*n, %*) |  |
| Left hippocampus |  |  |  |  |  |  |  |
| (%CorrAgeSex) | 0.190 | 0.033 | 9, 11 | 0.204 | 0.025 | 1, 2.5 | *U* = 1232, *P* = 0.04 |
| (cm<sup>3</sup>) | 2.075 | 0.427 | 19, 23.2 | 2.270 | 0.233 | 0 | - |
| Right hippocampus |  |  |  |  |  |  |  |
| (%CorrAgeSex) | 0.188 | 0.034 | 4, 4.9 | 0.201 | 0.031 | 0 | *U* = 1311, *P* = 0.09 |
| (cm<sup>3</sup>) | 2.058 | 0.450 | 10, 12.2 | 2.221 | 0.292 | 1, 2.5 | - |
| Left thalamus |  |  |  |  |  |  |  |
| (%CorrAgeSex) | 0.642 | 0.068 | 14, 17.1 | 0.686 | 0.055 | 1, 2.5 | *U* = 1002, *P* = 0.002 |
| (cm<sup>3</sup>) | 7.021 | 0.915 | 6, 7.3 | 7.609 | 0.934 | 0 | - |
| Right thalamus |  |  |  |  |  |  |  |
| (%CorrAgeSex) | 0.638 | 0.069 | 7, 8.5 | 0.665 | 0.062 | 1, 2.5 | *U* = 1228, *P* = 0.04 |
| (cm<sup>3</sup>) | 6.986 | 0.938 | 6, 7.3 | 7.362 | 0.934 | 0 | - |
| Left caudate |  |  |  |  |  |  |  |
| (%CorrAgeSex) | 0.371 | 0.054 | 5, 6.1 | 0.368 | 0.040 | 1, 2.5 | *U* = 1621, *P* = 0.46 |
| (cm<sup>3</sup>) | 4.054 | 0.634 | 3, 3.7 | 4.063 | 0.487 | 0 | - |
| Right caudate |  |  |  |  |  |  |  |
| (%CorrAgeSex) | 0.363 | 0.050 | 0 | 0.355 | 0.044 | 1, 2.5 | *U* = 1513, *P* = 0.39 |
| (cm<sup>3</sup>) | 3.963 | 0.579 | 2, 2.4 | 3.932 | 0.509 | 0 | - |
| Left putamen |  |  |  |  |  |  |  |
| (%CorrAgeSex) | 0.467 | 0.051 | 3, 3.7 | 0.464 | 0.043 | 0 | *U* = 1623, *P* = 0.46 |
| (cm<sup>3</sup>) | 5.091 | 0.493 | 1, 1.2 | 5.111 | 0.474 | 0 | - |
| Right putamen |  |  |  |  |  |  |  |
| (%CorrAgeSex) | 0.473 | 0.053 | 2, 2.4 | 0.469 | 0.044 | 0 | *U* = 1545, *P* = 0.39 |
| (cm<sup>3</sup>) | 5.164 | 0.539 | 0 | 5.176 | 0.551 | 1, 2.5 | - |
| Left hemisphere |  |  |  |  |  |  |  |
| (%CorrAgeSex) | 49.98 | 0.66 | 5, 6.1 | 49.95 | 0.50 | 0 | *U* = 1550, *P* = 0.39 |
| (cm<sup>3</sup>) | 550.3 | 56.7 | 1, 1.2 | 549.5 | 52.5 | 0 | - |
| Right hemisphere |  |  |  |  |  |  |  |
| (%CorrAgeSex) | 50.02 | 0.66 | 4, 4.9 | 50.05 | 0.50 | 0 | *U* = 1550, *P* = 0.39 |
| (cm<sup>3</sup>) | 550.2 | 56.5 | 1, 1.2 | 551.4 | 51.3 | 0 | - |

**Figure 2.**
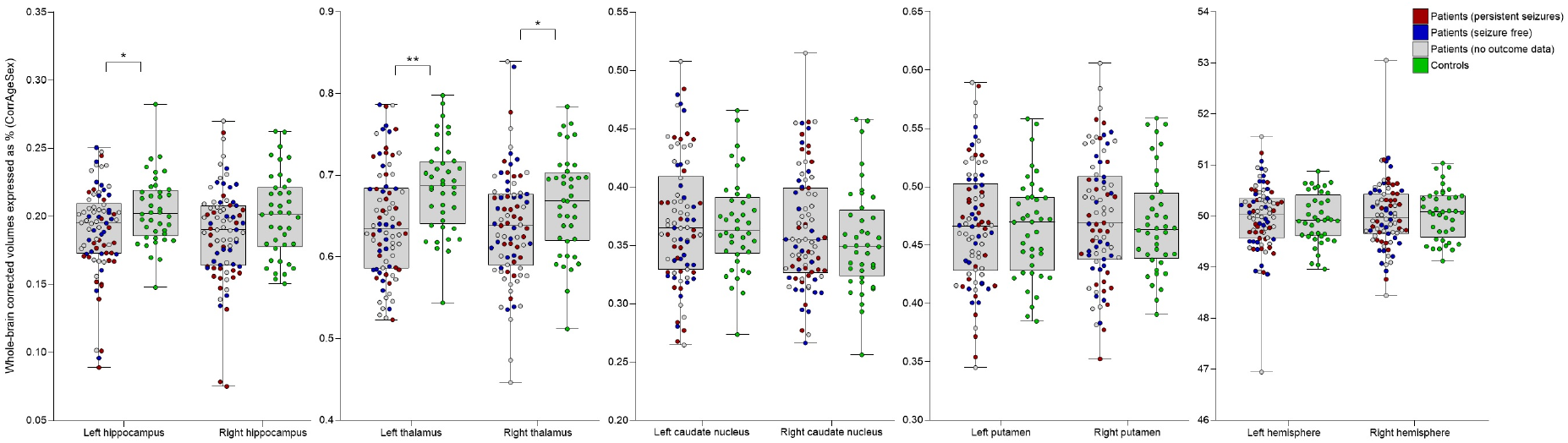
Scatterplots with minimum, 25th percentile, median, 75th percentile, and maximum normalised and corrected volume measurements of brain structures in seizure free patients, patients with persistent seizures, and healthy controls. Statistically significant between groups: *(*P*_(FDRcorr)_ < 0.05), **(*P*_(FDRcorr)_ < 0.01).

### Correlations with clinical variables

There were no associations between volumes and seizure outcome at 6 or 12 months, EEG finding, loss of awareness during seizures, history of FBTCS, age at first seizure, years between first seizure and diagnosis or age at diagnosis. Table 3 presents the descriptive and statistical comparisons between outcome groups at 12 months. There were also no clinically significant differences between the individual patients who had significant loss of hippocampal and thalamic volume and those who did not.

**Table 3.** Results of clinical and volumetric comparisons between seizure free patients and patients with persistent seizures at 12 months. FAS, focal aware seizures; FIAS, focal impaired awareness seizures; FBTCS, focal to bilateral tonic-clonic seizures; EEG, electroencephalography. †FBTCS data not available for one patient with persistent seizures. ‡EEG data not available for 11 (45.8%) seizure free patients and 10 (41.7%) patients with persistent seizures.

| Structure | Seizure free | | Persistent seizures | | $\chi^2$, $U$, $P(\text{FDRcorr})$ |
| --- | --- | --- | --- | --- | --- |
| $n$ (%) | 24 (50) | | 24 (50) | | - |
| Male / female | 14 (58.3) | 10 (41.7) | 14 (58.3) | 10 (41.7) | $\chi^2 = 0$, $P = 1.0$ |
| FAS / FIAS | 11 (45.8) | 13 (54.2) | 7 (29.2) | 17 (70.8) | $\chi^2 = 1.42$, $P = 0.49$ |
| FTBS / no FTBS† | 22 (91.7) | 2 (8.3) | 18 (75) | 5 (20.8) | $\chi^2 = 1.67$, $P = 0.49$ |
| Normal / abnormal EEG‡ | 11 (45.8) | 2 (8.3) | 7 (29.2) | 7 (29.2) | $\chi^2 = 3.64$, $P = 0.41$ |
| Mean / SD |  |  |  |  |  |
| Age | 39.71 | 12.22 | 37.00 | 10.25 | $U = 251$, $P = 0.45$ |
| Age of onset | 32.21 | 12.33 | 29.33 | 11.01 | $U = 250$, $P = 0.45$ |
| Left hippocampus |  |  |  |  |  |
| (%CorrAgeSex) | 0.193 | 0.032 | 0.175 | 0.035 | $U = 185$, $P = 0.34$ |
| (cm<sup>3</sup>) | 2.101 | 0.369 | 1.926 | 0.507 | - |
| Right hippocampus |  |  |  |  |  |
| (%CorrAgeSex) | 0.189 | 0.028 | 0.172 | 0.041 | $U = 201$, $P = 0.37$ |
| (cm<sup>3</sup>) | 2.070 | 0.336 | 1.892 | 0.520 | - |
| Left thalamus |  |  |  |  |  |
| (%CorrAgeSex) | 0.651 | 0.071 | 0.653 | 0.068 | $U = 280$, $P = 0.98$ |
| (cm<sup>3</sup>) | 7.156 | 0.906 | 7.157 | 0.917 | - |
| Right thalamus |  |  |  |  |  |
| (%CorrAgeSex) | 0.654 | 0.064 | 0.646 | 0.051 | $U = 257$, $P = 0.98$ |
| (cm<sup>3</sup>) | 7.209 | 0.923 | 7.084 | 0.792 | - |
| Left caudate |  |  |  |  |  |
| (%CorrAgeSex) | 0.367 | 0.053 | 0.365 | 0.058 | $U = 288$, $P = 1.0$ |
| (cm<sup>3</sup>) | 4.038 | 0.647 | 3.982 | 0.577 | - |
| Right caudate |  |  |  |  |  |
| (%CorrAgeSex) | 0.355 | 0.052 | 0.363 | 0.051 | $U = 255$, $P = 0.98$ |
| (cm<sup>3</sup>) | 3.894 | 0.614 | 3.963 | 0.503 | - |
| Left putamen |  |  |  |  |  |
| (%CorrAgeSex) | 0.468 | 0.045 | 0.462 | 0.055 | $U = 276$, $P = 0.98$ |
| (cm<sup>3</sup>) | 5.134 | 0.367 | 5.059 | 0.618 | - |
| Right putamen |  |  |  |  |  |
| (%CorrAgeSex) | 0.469 | 0.047 | 0.461 | 0.054 | $U = 271$, $P = 0.98$ |
| (cm<sup>3</sup>) | 5.150 | 0.528 | 5.042 | 0.571 | - |
| Left hemisphere |  |  |  |  |  |
| (%CorrAgeSex) | 50.00 | 0.63 | 49.94 | 0.54 | $U = 264$, $P = 0.98$ |
| (cm<sup>3</sup>) | 553.7 | 56.2 | 549.7 | 56.7 | - |
| Right hemisphere |  |  |  |  |  |
| (%CorrAgeSex) | 50.00 | 0.63 | 50.06 | 0.54 | $U = 264$, $P = 0.98$ |
| (cm<sup>3</sup>) | 553.1 | 55.4 | 550.8 | 56.6 | - |

## Discussion

In the present study we sought to establish whether structures that are known to show atrophy in focal epilepsy also show evidence of volume loss at the time of diagnosis of epilepsy. We report significant volume loss of the thalamus and hippocampus in adults with NDfE. Additionally, we aimed to explore whether clinical variables were associated with volume changes. Atrophy of subcortical structures was not related to seizure outcome at either 6 or 12 months or any other clinical characteristic of epilepsy.

### Biological and clinical implications

Of the limited number of quantitative MRI studies that exist in NDfE, the focus has been on the hippocampus. Our results are in support of those studies that reported hippocampal atrophy at diagnosis ^[14, 15, 26]^ and are in contrast to those that did not report atrophy ^[16]^. Patients with chronic epilepsy have shown a 13 to 16% reduction in hippocampal volume compared to controls ^[26]^; we identified a 6.5 to 6.9% decrease in hippocampal volume which may indicate hippocampal atrophy is present at time of diagnosis and may worsen with the progression of epilepsy. The present study also sought to determine whether extrahippocampal subcortical atrophy was present at diagnosis. To our knowledge, the present study is the first to identify thalamic atrophy in adults with a new diagnosis of focal epilepsy. A previous study in a small sample of patients with NDfE did not report thalamic atrophy ^[16]^. Thalamic atrophy has been reported to be almost as common as hippocampal atrophy in a meta-analysis of voxel-based morphometry studies of refractory TLE ^[27]^, and is observed in a range of longstanding focal and generalised epilepsy disorders ^[2, 28]^. Thalamic volume loss has also been reported in children with NDfE ^[29]^ and patients with new-onset GGE ^[19]^. Consistent with our findings, volume loss of the thalamus in both hemispheres is frequently reported in patients with chronic focal epilepsy ^[2, 28, 30-32]^. Taken together, our results suggest that thalamohippocampal atrophy is likely established prior to the onset of habitual epilepsy; further hippocampal and thalamic damage may occur as the disorder becomes longstanding, particularly in refractory cases ^[33-35]^.

There are very few existing studies that have attempted to predict pharmacoresistance from the point of diagnosis of focal epilepsy using advanced imaging in a way that resembles work predicting surgical outcome in focal epilepsy ^[31, 36]^. This is an unmet need in the early stages of the disorder ^[7, 8, 37]^. Having a reliable imaging biomarker of the health issues patients will experience (e.g. uncontrolled seizures, memory impairment) from diagnosis will provide clinicians and patients with realistic expectations and could serve to assist the patient management pathway (e.g. earlier use of adjunctive / alternative therapies in patients likely to be pharmacoresistant) ^[37]^. In the present study we have reported that gross neuroanatomical volume of subcortical structures are not related to seizure control at 6 or 12 months after diagnosis. (We note the relatively brief follow up; however, 12-month outcome is highly predictive of pharmacoresistance in NDE ^[12]^.) The advantage of investigating potential imaging biomarkers using MRI acquired as part of standard diagnostic evaluation is the applicability of findings to routine clinical practice. However, the disadvantage is that highly variable gross brain morphology is unlikely to be sensitive enough to identify markers of pharmacoresistance. It is likely that these markers, if found, will be microstructural, functional, or metabolic. Interestingly, in patients with longstanding focal epilepsy, MR spectroscopy hippocampal *N*-Acetylaspartate/Creatine measurements have been related to seizure control ^[38]^.

Approximately 50% of patients with NDfE exhibit impairment in at least one cognitive domain ^[13]^. Although we did not assess cognition in the current study, atrophy of subcortical structures may be related to cognitive impairment in patients with NDfE. Previous research has shown hippocampal volume loss is associated with poorer verbal memory performance in NDfE ^[14]^ and higher seizure frequency after AED withdrawal in TLE ^[39]^. Lower thalamic volume and higher seizure frequency have been observed in cognitively impaired children with NDE ^[29]^. Future research of imaging and cognitive biomarkers of pharmacoresistance in NDfE would be beneficial in establishing the most effective treatment pathways for patients likely to experience refractory epilepsy ^[37]^.

### Methodological issues

The discrepancy between our findings and those of a previous study that did not report thalamic atrophy in patients with NDfE ^[16]^ is difficult to reconcile. We have previously demonstrated good agreement between stereology and Freesurfer analysis – the method used by the previous study – for volume estimation of the thalamus ^[23]^; therefore image analysis approach is unlikely to be a factor. The substantially larger sample size of the present study is also unlikely to explain this discrepancy as the previous study reported a small trend for *increased* thalamic volume in patients with NDfE ^[16]^. Both studies investigated only patients with non-lesional NDfE. The only remaining difference between studies is type of MRI scans. The previous study used conventional three-dimensional (3D) T1-weighted scans with isotropic voxels (1 mm × 1 mm × 1mm), which offers limited differentiation between thalamic grey matter and adjacent white matter ^[23]^. We used non-isotropic T1-weighted FLAIR with high resolution in-plane coronal sections (0.4 mm × 0.4 mm), which provides superior delineation between the thalamus and white matter (Figure 1).

The standard MRI protocol for patients with a new presentation of seizures at our institution does not include 3D volume scans that would be amenable to automated segmentation techniques. It was necessary for us to apply manual volumetric analysis due to the 3 mm slice thickness of the coronal 2D T1-weighted FLAIR MRI scans. Despite this being a time-inefficient way of obtaining morphometric data, there were distinct advantages to our approach. Firstly, manual measurement of brain regions is considered gold standard and stereology provides a mathematically unbiased and validated approach to estimate brain compartment volume ^[21, 25]^. Secondly, despite the non-isotropic voxel size, the high in-plane resolution and contrast of the scans provided excellent grey-white matter differentiation, even in regions where the grey matter and white matter borders are difficult to establish (e.g. the thalamus).

Following diagnostic MRI most patients with NDfE will not undergo further investigation, which is usually undertaken in those with a refractory course, for who more precise localisation of the seizure focus is more likely as more seizures are witnessed and following increasingly detailed imaging, EEG and neuropsychological evaluation. Our sample of patients with NDfE is therefore likely to be clinically heterogenous in terms of seizure foci, despite commonalities in a new diagnosis of focal epilepsy and non-lesional MRI. We suggest that what is lost through the inclusion of a highly phenotyped group of patients is gained through a pragmatic approach to studying all non-lesional patients with a new diagnosis of focal epilepsy. Indeed, studying patients with NDfE pragmatically may yield common brain abnormalities, and potentially biomarkers of treatment outcome, given that; (i) widespread alterations in brain network structure can give rise to a clearly localised focal onset in one brain region ^[40]^; (ii) particular anatomical circuits act as critical modulators of seizure generation and propagation, and seizure activity does not spread diffusely throughout the brain but propagates along specific anatomical pathways, regardless of the localisation of the brain insult ^[41, 42]^; (iii) pathological structural connectivity causes disturbances to common large scale functional brain networks regardless of the localisation of the epileptogenic zone in patients with refractory focal epilepsy ^[43]^; and (iv) subcortical structures - such as the thalamus and striatum - that play a crucial role in the clinical manifestation of seizures in *the epilepsies* ^[44]^, and anatomically support widespread distributed cortico-subcortical networks ^[45]^, are structurally and physiologically abnormal in both hemispheres in patients with longstanding focal and generalised epilepsy disorders ^[2, 28, 44]^.

## Conclusions

Many specialist institutions and research centres do not see patients with epilepsy until it is well established, which may contribute to the lack of imaging studies of NDfE. In the present imaging study, we have studied a comparatively large number of patients with NDfE and report that atrophy of the hippocampi and thalami – ordinarily reported to be atrophic in longstanding and refractory focal epilepsy – is established at the time of diagnosis. It remains uncertain as to whether this atrophy is congenital, a consequence of epileptogenic processes or a combination of both.

## Data Availability

The data that support the findings of this study are available from the corresponding author, SSK, upon reasonable request.

## Acknowledgements

This work was funded by UK Medical Research Council (MR/S00355X/1 and MR/K023152/1) and Epilepsy Research UK (grant number 1085) grants awarded to SSK.

